# Is Left Atrial Appendage Closure a Universal Alternative to NOACs? A Meta-Analysis of NOAC-Era Trials

**DOI:** 10.64898/2026.05.24.26353968

**Authors:** Muhammad A. Bodla, Muhammad A. Mustehsan, Muhammad M. Shehzad, Saira Afzal

**Author notes:** These authors contributed equally to this work (Muhammad A. Bodla, Muhammad A. Mustehsan). Address for Correspondence: Muhammad A. Bodla, MBBS University College of Medicine & Dentistry, University of Lahore 1-Km Defence Road, Lahore, Punjab, Pakistan X (formerly Twitter): @AwaisBodla6.

## Abstract

**Background:** Non-vitamin K antagonist oral anticoagulants (NOACs) are the guideline-recommended standard for stroke prevention in atrial fibrillation (AF), yet bleeding risks limit real-world adherence. Percutaneous left atrial appendage closure (LAAC) offers a mechanical alternative without definitive comparative synthesis.

**Objectives:** To evaluate percutaneous LAAC versus NOAC therapy by synthesizing all contemporary NOAC-era randomized controlled trials (RCTs).

**Methods:** Five databases and registries (PubMed, MEDLINE, Embase, Cochrane CENTRAL, ClinicalTrials.gov) were searched from inception to 8 May 2026 for RCTs comparing percutaneous LAAC against NOACs in adults with non-valvular AF. Risk of bias was assessed using Cochrane RoB 2. Ischemic stroke was pooled using a random-effects DerSimonian-Laird model; primary efficacy composite and non-procedural bleeding were evaluated via pre-specified narrative synthesis.

**Results:** Four RCTs (CHAMPION-AF, OPTION, PRAGUE-17, CLOSURE-AF) comprising 5,890 patients were included. LAAC achieved noninferiority for the primary efficacy composite in three trials and demonstrated a statistically significant 45–56% reduction in non-procedural bleeding across the three moderate-risk trials. CLOSURE-AF did not meet noninferiority but retained a directionally consistent bleeding reduction. Pooled ischemic stroke analysis (HR 1.31; 95% CI 0.96–1.80; I²=0%) showed no statistically significant increase in stroke risk, though a consistent directional trend toward more ischemic events was observed.

**Conclusions:** LAAC significantly reduces non-procedural bleeding in moderate-risk AF patients, though this benefit attenuates in very high-risk populations. A consistent, statistically nonsignificant ischemic stroke trend and population-dependent efficacy establish LAAC as a shared decision-making alternative to NOACs rather than a universal replacement, pending 5-year CHAMPION-AF data.

**CONDENSED ABSTRACT:** This meta-analysis of four randomized controlled trials (5,890 patients) compared percutaneous left atrial appendage closure (LAAC) with NOAC therapy in non-valvular atrial fibrillation. LAAC significantly reduced non-procedural bleeding by 45–56% in moderate-risk patients, though this benefit attenuated in very high-risk, elderly cohorts. While successfully freeing eligible patients from lifelong anticoagulation, LAAC was associated with a consistent but statistically nonsignificant trend toward increased ischemic stroke. These clinical trade-offs establish LAAC as a highly effective, shared decision-making alternative to NOACs for carefully selected patients, emphasizing individualized risk profiling over universal device eligibility.

## INTRODUCTION

Atrial fibrillation (AF) is the most prevalent sustained cardiac arrhythmia worldwide, conferring an approximately five-fold increase in thromboembolic stroke risk and a two-fold increase in mortality^1^. Non-vitamin K antagonist oral anticoagulants (NOACs) have effectively replaced vitamin K antagonists as the guideline-recommended standard of care for stroke prevention across age groups in patients with a CHA DS -VASc score of at least 2^2^. However, managing high-risk patients remains challenging, as the real-world efficacy of NOACs is frequently limited by medication non-compliance and inherent bleeding risks^3,4^

Percutaneous left atrial appendage closure (LAAC) offers a mechanical alternative to lifelong anticoagulation by sealing the primary anatomical source of AF-related thrombi ^5^. Over the past decade, LAAC technology has matured significantly, transitioning from early-generation devices to contemporary iterations such as WATCHMAN FLX and AMPLATZER Amulet.^6,7^ This evolution has dramatically improved procedural safety; early devices carried complication rates near 8%, whereas contemporary large-scale registry data confirm that real-world implant success rates now exceed 97% with major complication rates below 3% in routine practice.^8,9^

Driven by this improved safety profile, the clinical paradigm is shifting from viewing LAAC solely as a backup strategy for “OAC failure” to positioning it as a first-line “OAC alternative” in eligible patients. ^10^ While multiple prior meta-analyses have evaluated LAAC, the most methodologically rigorous recent synthesis included only two trials with a NOAC comparator and predated the publication of CHAMPION-AF and CLOSURE-AF. ^11–17^

The present meta-analysis is the first to pool data exclusively from contemporary NOAC-era randomized controlled trials (RCTs) directly comparing LAAC against NOAC therapy. By nearly doubling the analyzed evidence base to 5,890 patients, we aim to definitively evaluate the comparative efficacy and safety of these two stroke prevention strategies, with a focus on pooling ischemic stroke as the primary quantitative outcome and providing narrative syntheses for primary efficacy and non-procedural bleeding. ^17^

## METHODS

### Protocol and Registration

This systematic review and meta-analysis was conducted in accordance with PRISMA 2020 guidelines. The protocol was registered on PROSPERO (CRD420261382375).

### Eligibility Criteria

Studies were eligible if they were prospective RCTs enrolling adults aged 18 years or older with non-valvular AF and a CHA DS -VASc score of at least 2 in men or at least 3 in women, comparing percutaneous LAAC against NOAC therapy, with at least one pre-specified outcome and a minimum follow-up of 12 months. Studies using warfarin as the sole comparator were excluded.

### Information Sources and Search Strategy

A systematic search of PubMed (n=222), Cochrane CENTRAL (n=129), Medline (via Ovid; n=204), and Embase (via Ovid; n=123) was conducted from inception to 8 May 2026, with no language restrictions. Additionally, the ClinicalTrials.gov registry (n=26) was searched to identify ongoing or recently completed trials. Search terms combined MeSH descriptors, Emtree terms, and free-text terms for atrial fibrillation, left atrial appendage closure, and anticoagulants. Full search strings are provided in the supplementary appendix. Duplicate records were identified and removed using Rayyan^18^ prior to screening. Ethical approval and patient consent were not required for this study, as it is a systematic review and meta-analysis of previously published randomized controlled trials. All analyzed data were derived from de-identified, aggregated results of studies that had already obtained their respective institutional review board approvals.

### Risk of Bias Assessment

Risk of bias was assessed using the Cochrane RoB 2 tool, independently by two reviewers, across five domains: randomisation process, deviations from intended interventions, missing outcome data, measurement of outcomes, and selection of reported results. Because device-versus-drug comparisons are inherently open-label, the presence of blinded endpoint adjudication was specifically assessed as a mitigating factor within Domain 4.

### Statistical Analysis

Ischemic stroke was selected as the primary pooled quantitative efficacy outcome. Pooling of hazard ratios (HRs), subdistribution hazard ratios (sHRs), and incidence rate ratios (IRRs) was considered methodologically appropriate for stroke given that cumulative stroke incidences across all included trials ranged from 1.2% to 3.6%, well below the 0.2 cumulative incidence threshold established by Thom et al. above which differences between effect measures may introduce meaningful bias.^19^ Under rare event conditions, HR, sHR, and IRR converge mathematically toward a common relative risk estimate, as formally demonstrated by Tanaka et al., rendering their pooling on the log-hazard scale defensible.^20^ Heterogeneity across pooled stroke estimates was absent in all analyses (I²=0%), providing empirical support for the validity of this approach. A random-effects DerSimonian-Laird model was used to account for residual between-study variability in patient populations, follow-up durations, and comparator regimens. All analyses were performed in R (version 4.3.2) using the meta package. Outcome analyses were based on complete case data as reported by the included trials; no missing data imputation was required or performed.

Primary efficacy composite outcomes are presented as narrative synthesis rather than a pooled estimate for two pre-specified reasons: composite endpoint definitions differ across trials (CLOSURE-AF and PRAGUE-17 incorporated bleeding or procedural complications within their primary composites, unlike CHAMPION-AF and OPTION), and moderate heterogeneity (I²=42%) is attributable to population-level differences.

Non-procedural bleeding is similarly presented as narrative synthesis. Cumulative bleeding incidences (8.5% to 19.0%) exceed the 0.2 rare-event threshold established by Thom et al. above which pooling of different effect measures may introduce bias.^19^ Additionally, the proportional hazards assumption was violated for major bleeding in CLOSURE-AF, necessitating restricted mean survival time analysis in that trial, rendering its bleeding estimate incommensurable with hazard ratios from the other three trials.

For CLOSURE-AF, which did not report hazard ratios, IRRs were approximated from reported event counts and patient-years of follow-up using the Poisson method with 95% CIs. For OPTION, which did not report per-arm patient-years specifically for the stroke endpoint, the stroke IRR was calculated from raw event counts alone (9 device-group versus 10 anticoagulation-group events) using headcount denominators rather than person-time; this is acknowledged as a methodological limitation and the resulting wide confidence interval (0.36–2.20) reflects the small event numbers rather than imprecision introduced by the denominator approach. PRAGUE-17 reported sHRs using the Fine-Gray competing risks model. For the bleeding outcome, the CLOSURE-AF total major bleeding figure was corrected from the published aggregate to a non-procedural-only estimate (52 device-group versus 61 medical-therapy events), excluding 18 procedure-related bleeding events from the device arm as reported in CLOSURE-AF Table 3 of Landmesser et al.^21^ This correction was pre-specified in the analysis plan to ensure comparability with the non-procedural bleeding endpoints reported in CHAMPION-AF, OPTION, and PRAGUE-17. Since bleeding is presented as narrative synthesis rather than a formally pooled primary outcome, the impact of this correction on the primary analytical conclusions is limited; however, it is applied to ensure internal consistency of the comparative narrative across trials. Pre-specified leave-one-out sensitivity analyses were conducted by sequentially excluding each trial to assess the influence of individual studies on the pooled stroke estimate. Given that I²=0% across all four trials, these analyses do not investigate heterogeneity, which is absent, but rather quantify the contribution of each trial to the pooled HR and identify whether any single trial disproportionately drives the directional signal.

Although primary efficacy composite and non-procedural bleeding outcomes are presented as narrative syntheses for the reasons described above, exploratory pooled estimates are additionally reported for both outcomes in the supplementary figures. These exploratory analyses serve a specific methodological purpose: they function as internal consistency checks, demonstrating that even when different effect measures are forced onto a common log-hazard scale under the most optimistic rare-event assumptions, the resulting pooled estimates are directionally and numerically consistent with the narrative synthesis. For bleeding, the exploratory pooled HR of 0.58 corroborates the narrative description of 45–56% reductions across three trials. For the efficacy composite, the exploratory pooled HR of 1.07 is consistent with the narrative description of noninferiority in three of four trials. The analytical heterogeneity across trials, specifically CLOSURE-AF requiring restricted mean survival time for bleeding due to proportional hazards violation, PRAGUE-17 using Fine-Gray subdistribution HRs due to competing mortality risk, and CHAMPION-AF using standard Cox regression, reflects genuine differences in the underlying data structure of each trial rather than arbitrary analytical choices. The exploratory pooling therefore does not claim methodological equivalence of these measures but rather uses their convergence under rare-event conditions as evidence that the narrative synthesis is not sensitive to the choice of analytical framework.

## RESULTS

### Study Selection

Combined database (n=678) and register (n=26) searches yielded 704 records. After deduplication, 309 unique records were screened by title and abstract. Of these, 286 were excluded at the title and abstract stage as they did not meet pre-specified eligibility criteria. 23 records were retrieved for full-text review. At the full-text stage, 19 records were excluded with documented reasons given in Table S1 in supplementary appendix.

Four prospective randomized controlled trials (RCTs) involving a total of 5,890 patients met all eligibility criteria and were included in the quantitative and narrative synthesis: CHAMPION-AF, OPTION, PRAGUE-17, and CLOSURE-AF. A PRISMA 2020 flow diagram is provided in Figure 1.

**Figure 1.**
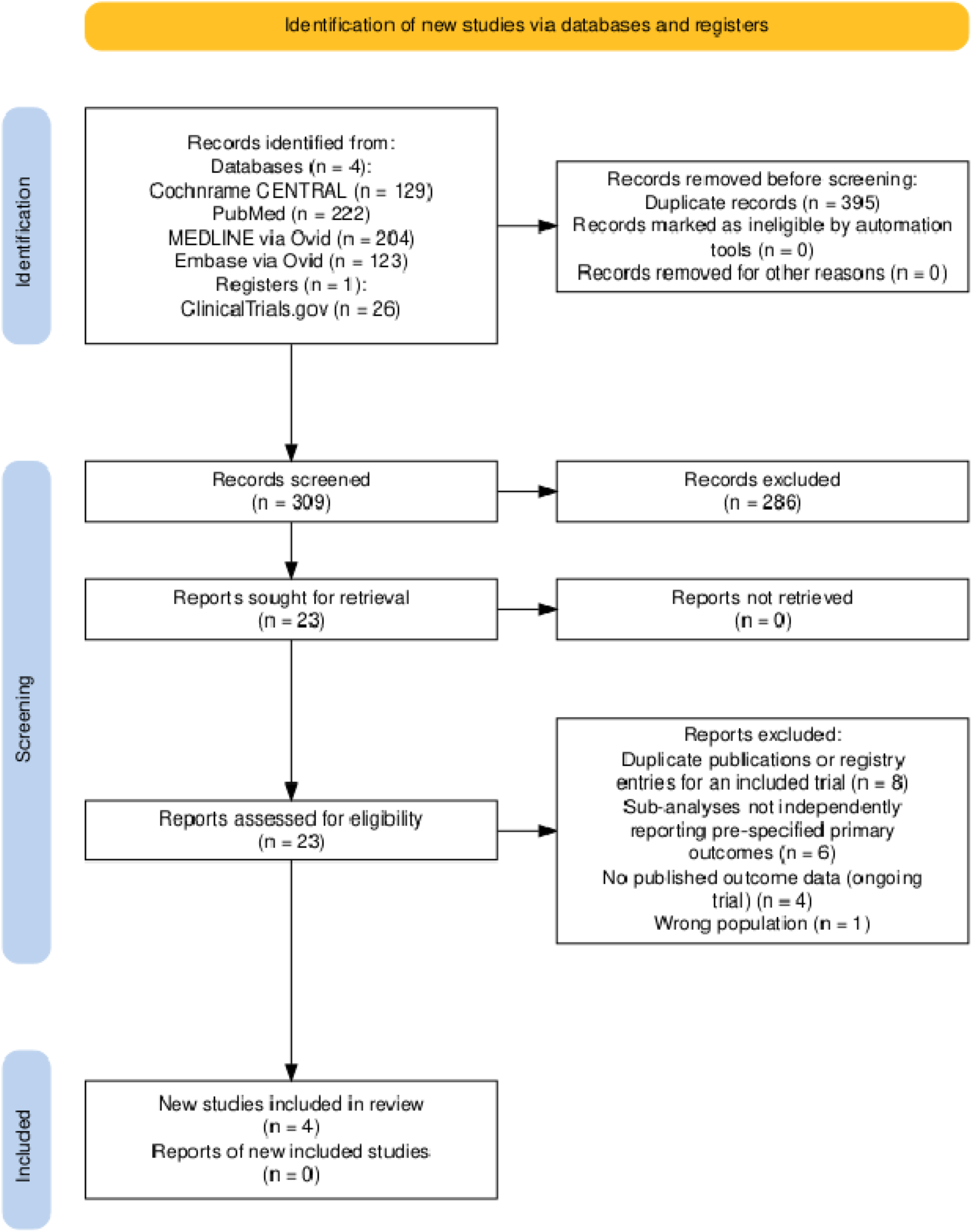
PRISMA 2020 Flow Diagram of Study Selection Systematic search and screening process from initial identification to final inclusion. Following the removal of 395 duplicates, 309 records were screened, resulting in the inclusion of 4 randomized controlled trials comprising 5,890 patients. PRISMA = Preferred Reporting Item for Systematic Reviews and Meta-Analyses.

### Study Characteristics

The four included trials comprised a total of 5,890 analysed patients (LAAC: 2,949; control: 2,941). CHAMPION-AF, a multicentre trial, randomized 3,000 moderate-risk patients (mean CHA DS -VASc 3.5) to receive the WATCHMAN FLX device versus NOAC therapy, evaluated over a 3-year follow-up.^22^ OPTION, also a multicentre trial, enrolled 1,600 moderate-risk patients (mean CHA DS -VASc 3.5) specifically following AF catheter ablation, comparing the WATCHMAN FLX device against NOAC therapy over 36 months.^23^ PRAGUE-17 randomized 402 high-risk patients (mean CHA DS -VASc 4.7) to a mix of LAAC devices (WATCHMAN, WATCHMAN FLX, or Amulet) versus DOAC therapy (predominantly apixaban) over a median follow-up of 3.5 years ^24,25^. CLOSURE-AF evaluated 888 very high-risk, older patients (mean CHA DS -VASc 5.2, mean age 78) randomized to mixed devices versus physician-directed best medical care (predominantly DOACs) over a median 3-year follow-up.^21^ Key baseline characteristics and individual trial estimates for all three primary outcomes are summarised in Tables 1 and 2.

**Table 1.**
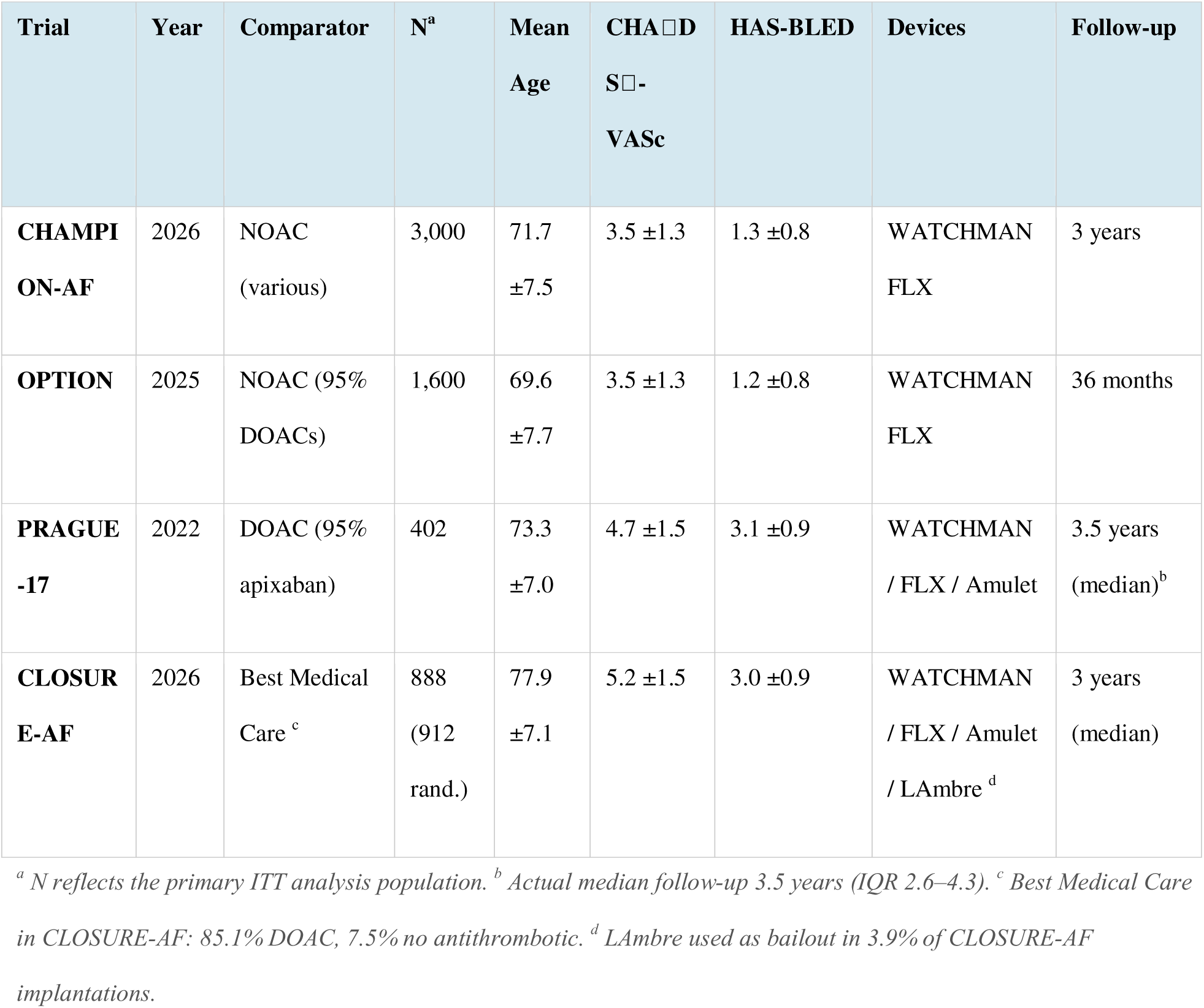
Baseline Characteristics and Design of Included Trials.

**Table 2.** Summary of Primary Outcomes by Trial.

| **Trial** | **Efficacy Composite**<br>**HR/sHR/IRR (95%**<br>**CI)□** | **Non-Procedural Bleeding**<br>**HR/sHR/IRR (95% CI)□** | **Stroke HR/sHR/IRR (95% CI)□** |
| --- | --- | --- | --- |
| **CHAMPION-**<br>**AF** | 1.20 (0.87–1.66) HR | 0.55 (0.45–0.67) HR <sup>b</sup> | 1.61 (1.00–2.59)<sup>c</sup> HR |
| **OPTION** | 0.91 (0.59–1.39) HR | 0.44 (0.33–0.59) HR <sup>b</sup> | 0.89 (0.36–2.20)<sup>d</sup> IRR |
| **PRAGUE-17** | 0.81 (0.56–1.18) sHR | 0.55 (0.31–0.97) sHR <sup>b</sup> | 1.14 (0.56–2.30)<sup>et</sup> sHR |
| **CLOSURE-AF** | 1.27 (1.00–1.60)<sup>g</sup> IRR | 0.89 (0.61–1.28)<sup>gh</sup> IRR | 1.20 (0.60–2.38)<sup>g</sup> IRR |

### Risk of Bias

A visual summary of the Cochrane RoB 2 assessment across all five domains for each included trial is presented in Supplementary Figure S1. All four trials are inherently open-label given the device-versus-drug comparison, which raises concerns in Domain 2 (deviations from intended interventions) and Domain 4 (measurement of outcomes). All four trials employed independent, blinded Clinical Events Committees for outcome adjudication as a Domain 4 mitigation: CLOSURE-AF adjudicated all events with committee members unaware of treatment assignment; PRAGUE-17 used a clinical endpoint committee blinded to patient allocation; OPTION and CHAMPION-AF both used independent Clinical Events Committees unaware of group assignments. OPTION, PRAGUE-17, and CLOSURE-AF are each rated as some concerns overall, reflecting the open-label design partially mitigated by blinded endpoint adjudication. CLOSURE-AF retains low risk in Domain 5 as no selective reporting concerns were identified, and low risk in Domain 3 given complete outcome data reporting. CHAMPION-AF is rated as high overall risk of bias due to two additional concerns independent of blinding: exclusion of procedural complications from the primary safety endpoint definition (Domain 5, selective reporting) and a 13.7% crossover from the NOAC arm to LAAC during the trial (Domain 2, deviations from intended interventions). Outside of the formal RoB 2 domains, it is notable that CLOSURE-AF and PRAGUE-17 are investigator-initiated and non-industry-funded, whereas CHAMPION-AF and OPTION are industry-funded.

### Primary Efficacy Composite: Narrative Synthesis

Noninferiority for the primary efficacy composite was met in CHAMPION-AF (HR 1.20, 95% CI 0.87–1.66)^22^, OPTION (HR 0.91, 95% CI 0.59–1.39)^23^, and PRAGUE-17 (sHR 0.81, 95% CI 0.56–1.18) ^24^. CLOSURE-AF, the only trial enrolling a very high-risk population (mean CHA DS -VASc 5.2, mean age 78 years, HAS-BLED 3.0), did not meet noninferiority (IRR 1.27, 95% CI 1.00–1.60)^21^. The pooled estimate across all four trials is HR 1.07 (95% CI 0.86–1.33, I²=42%); given composite definition heterogeneity and CLOSURE-AF’s population characteristics as a driver of heterogeneity, this figure is provided for reference only and not as the primary finding. Leave-one-out exclusion of CLOSURE-AF reduces heterogeneity to I²=24% and shifts the estimate to 0.98 (95% CI 0.77–1.25), consistent with noninferiority across the three moderate-risk trials. Exploratory pooled forest plots for the efficacy composite and leave-one-out sensitivity analyses are provided in Supplementary Figures S2, S4, S5, S6, and S7 alongside Supplementary Table S2. These exploratory analyses serve as internal consistency checks: even when HR, sHR, and IRR are treated as approximately equivalent under rare-event conditions, the resulting exploratory pooled estimate is numerically consistent with the narrative description of noninferiority in three of four trials, confirming that the clinical conclusion is not sensitive to the choice of analytical framework.

### Non-Procedural Bleeding: Narrative Synthesis

LAAC was associated with clinically meaningful reductions in non-procedural bleeding in CHAMPION-AF (HR 0.55, 95% CI 0.45–0.67)^22^, OPTION (HR 0.44, 95% CI 0.33–0.59)^23^, and PRAGUE-17 (sHR 0.55, 95% CI 0.31–0.97)^24^. These three trials, all enrolling moderate-risk patients (mean CHA DS -VASc 3.5–4.7), are highly consistent. In CLOSURE-AF^21^, restricting to non-procedural major bleeding events (52 device-group versus 61 medical-therapy events, excluding 18 procedure-related bleeds), the corrected IRR was 0.89 (95% CI 0.61–1.28), demonstrating a consistent directional trend.

Quantitative pooling of bleeding outcomes is not performed for the pre-specified reasons described in Statistical Analysis. The narrative pattern is clear: three of four trials demonstrate relative bleeding reductions of 45–56% with LAAC; the fourth (CLOSURE-AF) shows a corresponding directional trend, attenuated by high baseline bleeding risk, early procedure-related complications, and dual antiplatelet therapy requirements in a very high-risk elderly population. Individual trial estimates and pooled figures for all three outcomes are summarised in Table 2. Leave-one-out exclusion of CLOSURE-AF resolves bleeding heterogeneity completely (I² from 65% to 0%). Exploratory pooled forest plots for bleeding and leave-one-out sensitivity analyses are provided in Supplementary Figures S3, S8, S9, S10, and S11 alongside Supplementary Table S2. These exploratory analyses serve as internal consistency checks: even when HR, sHR, and IRR are treated as approximately equivalent under rare-event conditions, the exploratory pooled bleeding estimate of HR 0.58 (95% CI 0.44–0.76) is numerically consistent with the narrative description of 45–56% bleeding reductions across three trials, and the I² of 65% is fully explained by CLOSURE-AF’s distinct population, confirming that the narrative synthesis is not sensitive to the choice of analytical framework.

### Ischemic Stroke: Primary Pooled Analysis

Ischemic stroke was selected as the primary pooled quantitative efficacy outcome on the basis that it is the clinically central shared endpoint across all four trials, its cumulative incidences (1.2%–3.6%) satisfy the rare event conditions required for valid pooling of different effect measures, and it avoids the composite definition heterogeneity precluding direct pooling of primary efficacy endpoints.

A directional trend toward more ischemic strokes with LAAC was observed across three of four trials. In CHAMPION-AF^22^, the all-stroke HR was 1.46 (95% CI 0.94–2.27) and the ischemic stroke HR was 1.61 (95% CI 1.00–2.59). In PRAGUE-17^24^, the all-stroke/TIA sHR was 1.14 (95% CI 0.56–2.30). CLOSURE-AF^21^ demonstrated a corresponding directional trend (ischemic stroke IRR 1.20, 95% CI 0.60–2.38). OPTION^23^ (Wazni, 2025) showed a numerically lower rate in the device arm (IRR 0.89, 95% CI 0.36–2.20, from 9 device versus 10 NOAC events). The pooled HR was 1.31 (95% CI 0.96–1.80, P=0.09; I²=0%). Zero heterogeneity confirms this directional pattern is consistent across trials. The forest plot is presented in Figure 2.

**Figure 2.**
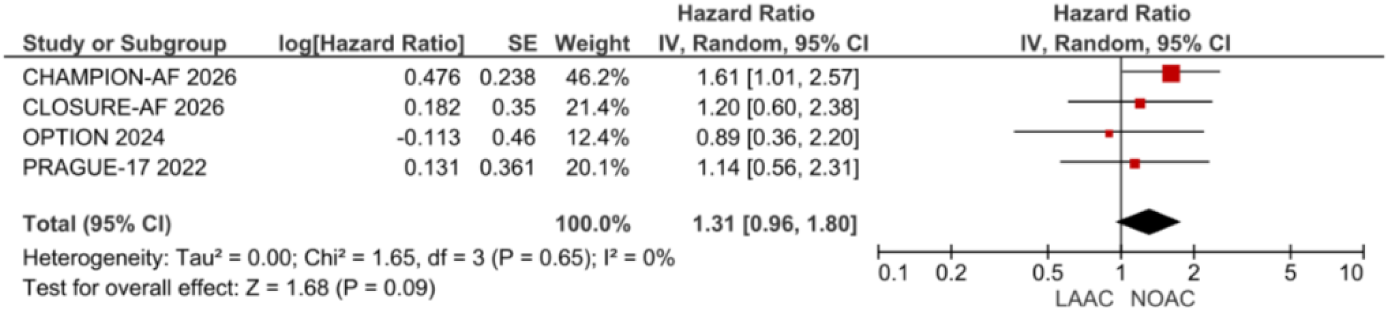
Ischemic Stroke: Primary Pooled Analysis (All Four Trials) This forest plot presents the primary pooled analysis of ischemic stroke using a random-effect DerSimonian-Laird model to evaluate the comparative efficacy of LAAC versus NOAC therapy. The pooled estimate demonstrates a corresponding directional trend toward more ischemic strokes with LAAC without between-trial heterogeneity, and pooling different effect measures on the log-hazard scale is mathematically justified under these rare-event conditions. CI = confidence interval; HR = hazard ratio; IRR = incidence rate ratio; LAAC = left atrial appendage closure; NOAC = non-vitamin K antagonist oral anticoagulant; sHR = subdistribution hazard ratio.

### Leave-One-Out Sensitivity Analyses: Ischemic Stroke

Critically, the directional trend toward more ischemic strokes with LAAC was preserved in every leave-one-out configuration without exception, with all four pooled estimates exceeding HR 1.0. The signal is therefore not dependent on any single trial but is a consistent finding across the evidence base. Pooled estimates ranged from HR 1.10 (95% CI 0.71–1.69, P=0.67) to HR 1.38 (95% CI 0.99–1.94), maintaining a corresponding directional trend. Heterogeneity remained absent in all configurations (I²=0%). CHAMPION-AF^22^, as the largest trial, contributes most to the precision of the estimate; however, its exclusion produced the most attenuated pooled estimate (HR 1.10) without reversing the direction, confirming the signal is not exclusively dependent on the largest trial. The leave-one-out analyses therefore demonstrate robustness of direction across all trial populations, from the post-ablation patients in OPTION^23^ to the very high-risk elderly patients in CLOSURE-AF^21^, strengthening rather than weakening confidence in the overall narrative. Numerical results are presented in Table 3. Leave-one-out forest plots are provided in Supplementary Figures S12, S13, S14, S15.

**Table 3.** Ischemic Stroke: Leave-One-Out Sensitivity Analyses.

| Analysis | Ischemic Stroke HR/sHR/IRR (95% CI) | I<sup>2</sup> | p |
| --- | --- | --- | --- |
| **All 4 Trials (primary)** | 1.31 (0.96–1.80) | 0.0% | 0.650 |
| **Excl. CLOSURE-AF** | 1.34 (0.93–1.92) | 0.0% | 0.457 |
| **Excl. CHAMPION-AF** | 1.10 (0.71–1.69) | 0.0% | 0.868 |
| **Excl. OPTION** | 1.38 (0.98–1.94) | 0.0% | 0.656 |
| **Excl. PRAGUE-17** | 1.36 (0.95–1.94) | 0.0% | 0.482 |

<sup>a</sup> Random-effects DerSimonian-Laird model. I<sup>2</sup>=0% across all configurations confirms no between-trial heterogeneity. Excluding CHAMPION-AF produces the most attenuated estimate (HR 1.10), confirming that the directional finding is robust to the exclusion of any single trial. Stroke metrics utilized: CHAMPION-AF = ischemic stroke HR; CLOSURE-AF = ischemic stroke IRR; OPTION = Poisson IRR (calculated from 9 vs 10 events); PRAGUE-17 = all-stroke/TIA sHR. For LOO analyses of the primary efficacy composite and non-procedural bleeding, see Supplementary Table S2 and Supplementary Figures S3–S10. CI = confidence interval; HR = hazard ratio; IRR = incidence rate ratio; LOO = leave-one-out; sHR = subdistribution hazard ratio; TIA = transient ischemic attack.

### Procedural Safety

Procedural complication rates have declined substantially across device generations: 8.0% in PROTECT-AF, 2.2% in PREVAIL^15^, 4.8% in PRAGUE-17 (9 of 187 patients)^24^, and 2.9% in OPTION (23 of 803)^23^, consistent with contemporary real-world data^9^ (Kapadia, 2024). Device-related thrombus was detected in 4.8% of CHAMPION-AF patients at 4 months, with 1.8% requiring anticoagulation resumption^22^. CLOSURE-AF reported two periprocedural deaths and five cases of pericardial tamponade among 446 LAAC patients. Ultimately, while overall complication rates reflect a maturing technology with high safety in moderate-risk patients, the adverse events in CLOSURE-AF underscore that very elderly, highly comorbid populations remain particularly vulnerable to upfront procedural risks.^21^

## DISCUSSION

### Novelty and Positioning

This meta-analysis is the first to pool all four NOAC-era RCTs comparing percutaneous LAAC directly against contemporary oral anticoagulation, nearly doubling the analysed evidence base from approximately 3,116 to 5,890 patients. Unlike recent network meta-analyses by Oliva et al.^26^ and Lerman et al.^27^, which relied on transitivity assumptions for indirect cross-trial comparisons, this analysis exclusively utilizes direct head-to-head RCT data. Furthermore, it supersedes the most methodologically rigorous immediate predecessor by Kaisaier et al.^17^, which reported a high-certainty relative risk of 1.38 for ischemic stroke or systemic embolism alongside a bleeding benefit, but was conducted without the definitive data from CHAMPION-AF and CLOSURE-AF.

### Primary Efficacy and Prior Literature

Before contextualizing the results of the present analysis, it is essential to ground the findings in the pre-2026 literature. Previous meta-analyses by Chen et al. (2021), Jiang et al. (2023), and Thapa et al. (2025) consistently identified reductions in bleeding and mortality with LAAC, while documenting a persistent directional trend toward increased ischemic stroke (e.g., an HR of ∼1.2).^28–30^ Therefore, the present analysis does not uncover a novel ischemic signal, but rather quantifies an existing one with higher-powered, contemporary data.

LAAC achieved noninferiority for the primary efficacy composite in CHAMPION-AF, OPTION, and PRAGUE-17. CLOSURE-AF, the trial enrolling the highest-risk population (mean CHA DS -VASc 5.2, mean age 78), did not meet noninferiority. This divergence is biologically coherent and directly supported by recent meta-regression analysis by Singh et al. (2025), which demonstrates that older, higher-risk patients experience an attenuated clinical benefit from LAAC.^31^ In lower-risk patients, the upfront procedural risk of LAAC is offset over time by the elimination of lifelong anticoagulation; conversely, in very high-risk, elderly patients, procedural complications accumulate before long-term benefits can materialize. However, to maintain scholarly balance, the critical perspective advanced by Saraf and Morris (2023) must be acknowledged, which argues based on early head-to-head data that LAAC may ultimately prove inferior to NOACs for ischemic stroke prevention.^32^

### Ischemic Stroke Risk and Antithrombotic Protocols

A directional trend toward more ischemic strokes with LAAC was observed across three of four trials, yielding a pooled HR of 1.31 (95% CI 0.96–1.80). The upper bound of the confidence interval (1.80) does not exclude a clinically meaningful increase in ischemic stroke risk, reinforcing the trend observed by Kaisaier et al.^17^ This residual ischemic risk highlights the clinical challenge of managing stroke despite oral anticoagulation, a phenotype for which LAAC is increasingly considered but where comparative data remain complex, as noted by Chatani et al. (2025).^33^

Device-related thrombus (DRT), detected in 4.8% of CHAMPION-AF patients, remains the most biologically plausible mechanism for this ischemic trend. Device-specific factors may influence this risk; for example, 5-year data from the Amulet IDE trial^34^ indicate that DRT and peridevice leak rates differ meaningfully between device generations (e.g., historically more frequent with WATCHMAN than Amulet), suggesting that future iterations may mitigate this hazard. Post-implant antithrombotic regimens also critically influence DRT. The ADALA trial^35^ demonstrated zero DRT cases with low-dose apixaban post-LAAC versus 8.7% with dual antiplatelet therapy.

Multiple ongoing trials, including comparisons of non-antithrombotic therapy versus single antiplatelet therapy ^36^ and low-dose rivaroxaban maintenance strategies, will test whether optimized, less-intensive post-procedural regimens can effectively suppress DRT and narrow the ischemic gap. Conversely, for patients who suffer ‘breakthrough’ ischemic strokes despite optimal anticoagulation the ongoing ELAPSE trial^37^ (formerly Occlusion-AF)^38^ will test the superiority of combination therapy (LAAC plus DOACs) against DOAC monotherapy. While these ongoing studies represent critical future directions, the pre-specified 5-year ischemic stroke endpoint from CHAMPION-AF will provide the definitive test of LAAC’s long-term stroke prevention efficacy.

### Non-Procedural Bleeding, ICH, and Surgical Context

In the present analysis, three of the four trials demonstrated relative non-procedural bleeding reductions of 45–56% with LAAC. This RCT-level bleeding dividend is robustly corroborated by a combined meta-analysis of randomized and propensity-matched studies, which identified a number needed to treat (NNT) of 25 for the prevention of major bleeding with LAAC compared to NOACs.^39^ Furthermore, these controlled trial findings translate directly to real-world populations, as validated by recent Medicare cohort data (n=26,979) demonstrating a substantial reduction in hospitalized bleeding among beneficiaries receiving LAAC compared to those on DOAC therapy.^40^

The clinical significance of this bleeding reduction is most profound when considering intracranial hemorrhage (ICH), the most feared consequence of oral anticoagulation, carrying an approximate 30-day mortality rate of 40% alongside severe long-term morbidity.^41^ For patients with a history of ICH or a high microbleed burden from cerebral amyloid angiopathy (CAA), systemic anticoagulation is essentially contraindicated^42^, even alternative pharmacological strategies utilizing antiplatelet agents carry nuanced hemorrhagic risks in this fragile cohort.^43^ By completely bypassing the pharmacological dilemma, LAAC provides a definitive mechanical solution. Large-scale observational evidence^44^ (Ajmal, 2020) and recent Bayesian network meta-analyses^45,46^ consistently rank LAAC as the optimal strategy for avoiding recurrent ICH in high-risk patients. Reflecting this paradigm shift, the World Stroke Organization recently issued a scientific statement supporting LAAC for secondary stroke prevention in patients for whom anticoagulation poses unacceptable risks.^47^ Consequently, the non-procedural bleeding reduction observed in this meta-analysis is clinically transformative for high-risk phenotypes, such as the prior-ICH patients enrolled in CLOSURE-AF.

The ability of LAAC to effectively decouple stroke prevention from systemic bleeding risk is anchored in strong biological plausibility. The mechanistic rationale for sealing the appendage is directly validated by the surgical LAAOS III trial. In that study, surgical LAA occlusion performed concomitantly with open-heart surgery reduced the risk of ischemic stroke or systemic embolism by an additional 33% even when patients continued full oral anticoagulation therapy.^48^ This surgical data provides definitive proof that the LAA is an independent, critical embolic source regardless of systemic antithrombotic therapy status, thereby validating the fundamental mechanistic concept underlying percutaneous LAAC and supporting its efficacy in the trials reviewed here.

### Special Populations and the Post-Ablation Context

The inclusion of the OPTION trial introduces a distinct post-ablation AF population into this synthesis. As demonstrated by a recent network meta-analysis of post-ablation antithrombotic strategies, ischemic stroke rates following successful catheter ablation are exceedingly low; approximately 0.23 per 100 person-years.^49^ In this specific, lower-risk context, demonstrating incremental stroke prevention efficacy is statistically challenging; consequently, the primary clinical objective for these patients shifts from stroke prevention to bleeding avoidance. This paradigm perfectly contextualizes the OPTION findings: for low-risk post-ablation patients, LAAC functions primarily as a highly effective bleeding-avoidance strategy, explaining why it was the only trial to show a numerically favorable ischemic trend for the device arm (IRR 0.89) while successfully demonstrating definitive superiority for non-procedural bleeding reduction.

### Procedural Safety and Concomitant Implantation

As procedural complication rates continue to decline with contemporary devices, the integration of LAAC into standard electrophysiology workflows has become increasingly viable. This is strongly supported by a recent subanalysis of the OPTION trial, which confirmed that concomitant, same-day LAAC and AF ablation procedures share a similar safety profile to staged, sequential procedures.^50^ These data alleviate historical concerns regarding combined-procedure risks and support the streamlining of patient care through single-setting interventions.

### Mortality in the NOAC Era

Unlike earlier meta-analyses comparing LAAC to warfarin ^15,16^, which demonstrated a significant cardiovascular mortality benefit for device therapy, the present analysis of NOAC-era trials does not replicate a mortality advantage. This shift is expected and robustly corroborated by a recent high-dimensional propensity score analysis of over 50,000 patients by Zhao et al. (2025).^51^ Their study confirmed that the mortality benefit of LAAC observed in older trials is extinguished when DOACs serve as the comparator. Because DOACs are intrinsically safer and carry a substantially lower risk of fatal intracranial hemorrhage than warfarin, the comparative mortality threshold is significantly higher today, perfectly aligning with the equivalent mortality rates observed in our synthesis.

### Future Directions

Looking forward, the evaluation of LAAC is expanding beyond traditional thromboembolic and bleeding endpoints. Emerging observational data from a large retrospective cohort study suggest that LAAC may be associated with a reduced incidence of dementia compared to oral anticoagulation. ^52^However, this remains a purely observational signal that requires rigorous prospective validation and was not an adjudicated endpoint in the trials reviewed here. Furthermore, optimizing antithrombotic management post-closure remains a critical frontier. The ongoing ATLAAC trial, which is definitively testing whether oral anticoagulation can be safely discontinued entirely following surgical LAAC^53^, will serve as an essential surgical companion to the percutaneous trials synthesized in this analysis, further clarifying the bounds of safe anticoagulation cessation.

### Heterogeneity

I² values of 42% for efficacy and 65% for bleeding are attributable to clinical rather than methodological sources, with CLOSURE-AF as the primary driver across both outcomes. This is expected given the stark population differences: CLOSURE-AF enrolled patients 8 to 9 years older on average, with substantially higher baseline stroke and bleeding risk scores, and utilized a mixed device and control arm strategy.^21^ Leave-one-out analyses formally confirm this: excluding CLOSURE-AF resolves bleeding I² to 0% and reduces efficacy I² to 24%. Conversely, the ischemic stroke I² of 0% across all analyses indicates a complete absence of between-trial heterogeneity for the primary pooled outcome. This is itself a finding worth highlighting: despite profound clinical divergence across the evidence base, ranging from the lower-risk post-ablation cohort in OPTION to the very high-risk elderly population in CLOSURE-AF, every trial produces a stroke estimate in the same direction, providing a strong statement about the consistency of this directional ischemic signal. Furthermore, the exploratory pooled estimates for efficacy (HR 1.07) and bleeding (HR 0.58) are numerically consistent with the narrative synthesis, confirming that analytical framework differences across trials do not meaningfully distort the clinical conclusions.

### Limitations

Only four trials were available for quantitative synthesis, substantially limiting statistical power. All risk-stratification observations should be treated as hypothesis-generating. Primary composite endpoints are not directly poolable due to composite definition differences. PRAGUE-17 reports sHRs rather than conventional Cox regression HRs; a pre-specified sensitivity analysis excluding PRAGUE-17 does not change directional conclusions. OPTION enrolled exclusively a post-catheter ablation population, limiting generalisability to broader non-valvular AF. Two of four trials (CHAMPION-AF and OPTION) are industry-funded. Furthermore, while CHAMPION-AF is rated at a high overall risk of bias, it is important to acknowledge that this rating is specifically domain-limited, driven by a 13.7% crossover rate from the control arm and the exclusion of procedural bleeding from the primary safety endpoint, rather than reflecting poor overall trial conduct or data quality. Finally, these findings derive from RCT populations with protocol-mandated DOAC adherence. Real-world DOAC discontinuation rates of 30–35%^54^ suggest that the observed bleeding reduction in these controlled trials may actually underestimate LAAC’s comparative “set-and-forget” advantage in routine clinical practice.

## CONCLUSIONS

This meta-analysis, the first to incorporate all four NOAC-era RCTs, demonstrates that percutaneous LAAC consistently reduces non-procedural bleeding in patients with moderate stroke and bleeding risk, but this benefit is attenuated in very high-risk, elderly patients. A directional trend toward more ischemic strokes with LAAC, consistent across three of four trials and confirmed by zero heterogeneity in leave-one-out analyses, requires close monitoring pending 5-year CHAMPION-AF data. These clinical trade-offs establish LAAC as a highly effective, shared decision-making alternative to NOACs in carefully selected patients with moderate CHA DS -VASc (2–4) and low-to-moderate HAS-BLED (<3) scores, successfully freeing eligible patients from lifelong anticoagulation. A visual summary of these comparative trial outcomes and our proposed patient selection framework is provided in the Central Illustration.

## CLINICAL PERSPECTIVES

### Competency in Medical Knowledge

Percutaneous left atrial appendage closure (LAAC) consistently reduces non-procedural bleeding by 45–56% in patients with moderate stroke and bleeding risk compared to NOACs, though this magnitude of benefit is attenuated in very high-risk, elderly populations.

### Competency in Patient Care

While LAAC successfully frees eligible patients from the burden of lifelong anticoagulation, it carries a consistent directional trend toward increased ischemic stroke. Clinicians must apply meticulous patient selection based on individual risk profiles, utilizing LAAC as a targeted alternative rather than a universal replacement for NOAC therapy.

### Competency in Interpersonal and Communication Skills

Given the clinical trade-offs between a significant reduction in non-procedural bleeding and a residual ischemic stroke risk, it is essential for clinicians to engage patients in shared decision-making to thoroughly discuss the distinct risks and benefits of device-based closure versus lifelong NOACs.

### Translational Outlook 1

The pre-specified 5-year ischemic stroke endpoint from the CHAMPION-AF trial represents the critical next step to definitively establish the long-term efficacy of LAAC as a first-line NOAC alternative and clarify the boundaries of residual ischemic risk.

### Translational Outlook 2

Further investigation into optimized, less-intensive post-implant antithrombotic protocols (such as those currently being evaluated in the ADALA^35^ and ANDES^55^ trials) is required to determine whether device-related thrombus rates can be effectively suppressed to narrow the observed ischemic gap

### Funding and Support

This research received no specific grant, contract, or other forms of financial support from any funding agency, foundation, fund, or institution in the public, commercial, or not-for-profit sectors.

### Clinical Trial Registration

The protocol for this systematic review and meta-analysis was registered on PROSPERO (CRD420261382375).

### Disclosures / Relationship with Industry

The authors have no relationships with industry or other relevant entities, financial or otherwise, to disclose. All authors are unfunded academic investigators.

### Proposed Tweet

Is LAAC the new standard over NOACs for AF? Our meta-analysis of 5,890 patients across contemporary RCTs shows LAAC reduces non-procedural bleeding by up to 56% in moderate-risk patients. A vital alternative for stroke prevention. #CardioTwitter #EPeeps #JACC @JACCJournals

## Data Availability

All data analyzed during this study are included in this published article and its supplementary information files. The underlying dataset consists entirely of aggregate-level data from clinical trial reports that are openly available in the public domain.

https://www.nejm.org/doi/abs/10.1056/NEJMoa2517213

https://www.nejm.org/doi/full/10.1056/NEJMoa2513310

https://www.nejm.org/doi/abs/10.1056/NEJMoa2408308

https://www.jacc.org/doi/10.1016/j.jacc.2020.04.06

## Abbreviations

AF: atrial fibrillation
CI: confidence interval
DOAC: direct oral anticoagulant
DRT: device-related thrombus
HR: hazard ratio
ICH: intracranial hemorrhage
IRR: incidence rate ratio
LAAC: left atrial appendage closure
NOAC: non-vitamin K antagonist oral anticoagulant
sHR: subdistribution hazard ratio

## Figures and captions

**CENTRAL ILLUSTRATION:**
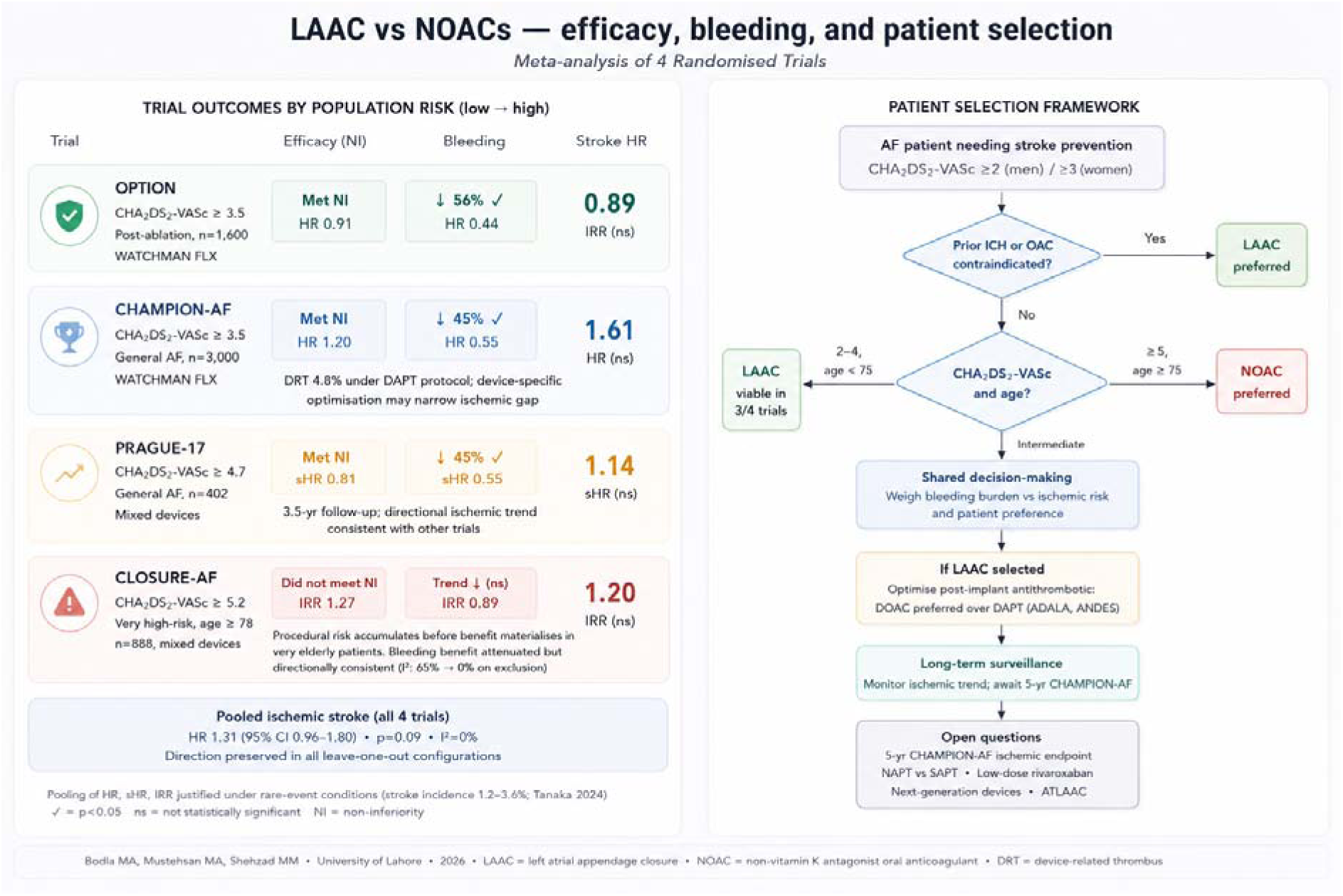
LAAC vs NOACs: efficacy, bleeding, and patient selection This illustration summarizes the outcomes and clinical application of LAAC versus NOACs in non-valvular atrial fibrillation. The left panel arrays four trials (OPTION, CHAMPION-AF, PRAGUE-17, CLOSURE-AF) by increasing baseline risk, demonstrating that LAAC’s efficacy non-inferiority and bleeding benefits in moderate-risk cohorts attenuate in highest-risk patients, alongside a directional trend toward increased ischemic stroke. The right panel translates these findings into a practical patient selection framework prioritizing shared decision-making. Abbreviations: AF = atrial fibrillation; CHA2DS2-VASc = congestive heart failure, hypertension, age ≥75 years, diabetes, prior stroke/transient ischemic attack, vascular disease, age 65-74 years, sex category; CI = confidence interval; DAPT = dual antiplatelet therapy; DRT = device-related thrombus; HR = hazard ratio; ICH = intracranial hemorrhage; IRR = incidence rate ratio; LAAC = left atrial appendage closure; NAPT = non-antithrombotic therapy; NI = non-inferiority; NOAC = non-vitamin K antagonist oral anticoagulant; OAC = oral anticoagulation; SAPT = single antiplatelet therapy; sHR = subdistribution hazard ratio.

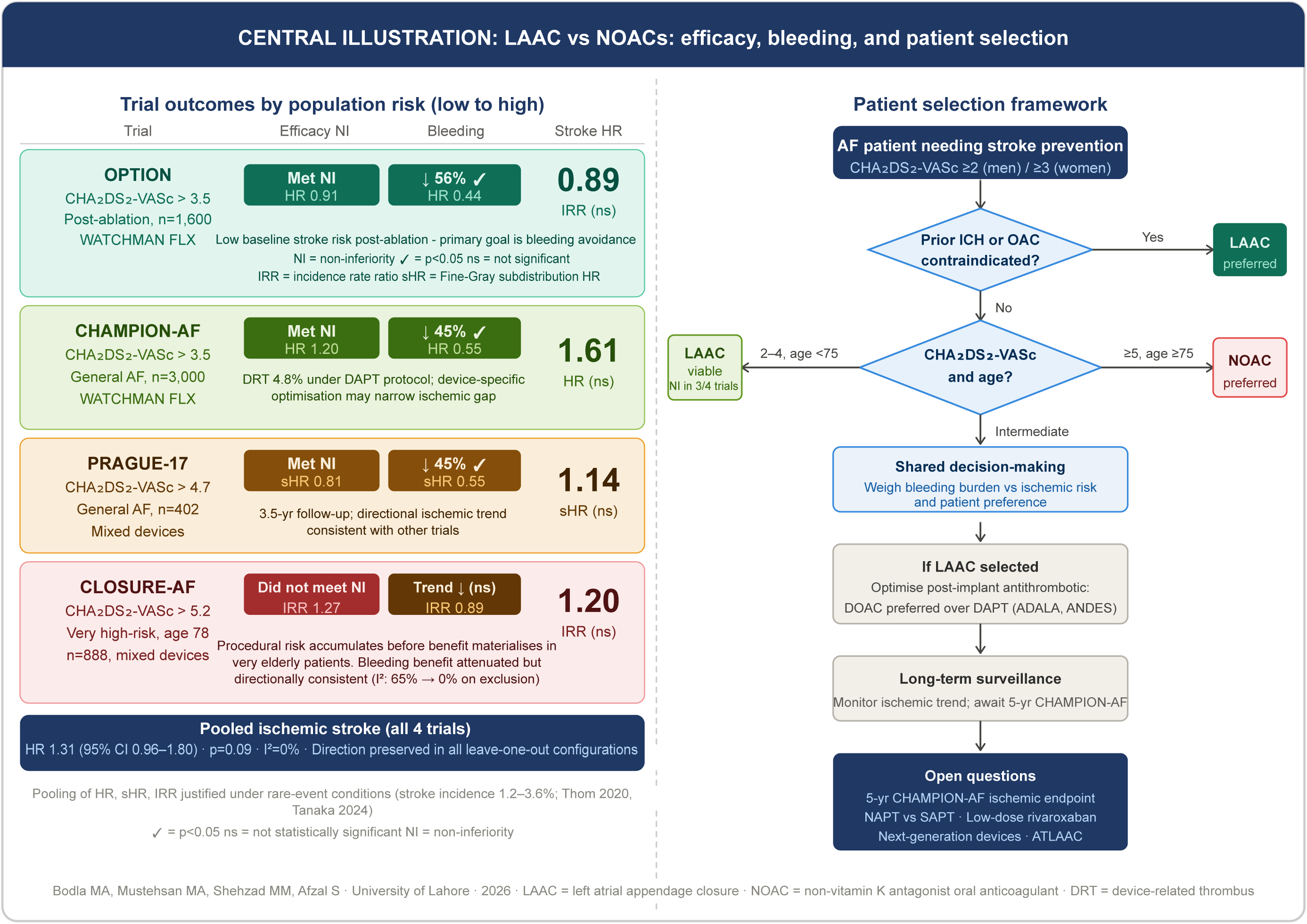

