## Supplemental Appendix for "Is Left Atrial Appendage Closure a Universal Alternative to NOACs? A Meta-Analysis of NOAC-Era Trials"

Page 1 - **Supplemental Methods:**

Page 1 - Search Strings

Page 6 - Full-Text Excluded Studies with Reasons

Page 9 - Risk of Bias Traffic Light Plot

Page 11 - **Supplemental Results:**

Page 11 - Exploratory pooled forest plots for the primary efficacy composite and non-procedural bleeding, provided as internal consistency checks for the narrative synthesis

Page 13 - Leave-one-out sensitivity analyses for all three outcomes. These analyses are provided for methodological transparency and are not primary findings. See Statistical Analysis in the main manuscript for full justification of the exploratory pooling approach.

### Supplemental Methods:

#### Search Strings

#### **Database 1: Cochrane Central Register of Controlled Trials (CENTRAL)**

| **Host** | Cochrane Library (https://www.cochranelibrary.com) |
| --- | --- |
| **Date searched** | 07 May 2026 at 21:04 |
| **Results** | 129 records |
| **RCT filter** | Not applied. CENTRAL indexes RCTs and controlled trials only by design |

#1	MeSH descriptor: [Atrial Fibrillation] explode all trees

#2	"atrial fibrillation":ti,ab,kw OR "non-valvular AF":ti,ab,kw

#3	#1 OR #2

#4	MeSH descriptor: [Left Atrial Appendage Closure] explode all trees

#5	(left atrial NEAR/3 appendage NEAR/3 (closur* OR occlusio* OR exclusio* OR ligatio*)):ti,ab,kw

#6	LAAC:ti,ab,kw OR LAAO:ti,ab,kw OR "WATCHMAN FLX":ti,ab,kw OR "Amplatzer Amulet":ti,ab,kw

#7	#4 OR #5 OR #6

#8	MeSH descriptor: [Anticoagulants] explode all trees

#9	(direct NEXT oral NEXT anticoagulant*):ti,ab,kw OR NOAC*:ti,ab,kw OR DOAC*:ti,ab,kw

#10 apixaban OR rivaroxaban OR dabigatran OR edoxaban

#11 #8 OR #9 OR #10

#12 #3 AND #7 AND #11	[129 results]

#### **Database 2: PubMed (MEDLINE + ahead-of-print)**

| **Host** | National Library of Medicine (https://pubmed.ncbi.nlm.nih.gov) |
| --- | --- |
| **Date searched** | 08 May 2026 |
| **Results** | 222 records |
| **RCT filter** | Cochrane-derived sensitivity/specificity balanced RCT filter applied |

((("Atrial Fibrillation"[Mesh] OR "atrial fibrillation"[tiab] OR "non-valvular AF"[tiab]))

AND

(("Left Atrial Appendage Closure"[Mesh]

OR "left atrial appendage closure"[tiab:~3]

OR "left atrial appendage occlusion"[tiab:~3]

OR "left atrial appendage exclusion"[tiab:~3]

OR "left atrial appendage ligation"[tiab:~3]

OR LAAC[tiab] OR LAAO[tiab]

OR "WATCHMAN FLX"[tiab]

OR "Amplatzer Amulet"[tiab]))

AND

(("Anticoagulants"[Mesh]

OR "direct oral anticoagulant*"[tiab]

OR NOAC*[tiab] OR DOAC*[tiab]

OR apixaban[tiab] OR rivaroxaban[tiab]

OR dabigatran[tiab] OR edoxaban[tiab])))

AND

((randomized controlled trial[pt]

OR controlled clinical trial[pt]

OR randomized[tiab]

OR placebo[tiab]

OR "clinical trials as topic"[mesh:noexp]

OR randomly[tiab]

OR trial[ti]

NOT (animals[mesh] NOT humans[mesh])))

[222 results]

#### **Database 3: MEDLINE via Ovid**

| **Host** | Ovid (https://ovidsp.ovid.com); MEDLINE(R) All, 1946 to present |
| --- | --- |
| **Date searched** | 08 May 2026 |
| **Results** | 204 records |
| **RCT filter** | Cochrane-derived sensitivity/specificity balanced RCT filter applied |
| **Note** | MEDLINE searched via Ovid in addition to PubMed to capture indexing differences between platforms. Duplicates with PubMed were removed electronically during deduplication. |

((exp Atrial Fibrillation/

or (atrial fibrillation or "non-valvular AF").ti,ab.)

AND

(exp Left Atrial Appendage Closure/

or (left atrial adj3 appendage adj3

(closur* or occlusio* or exclusio* or ligatio*)).ti,ab.

or (LAAC or LAAO or "WATCHMAN FLX" or "Amplatzer Amulet").ti,ab.)

AND

(exp Anticoagulants/

or ("direct oral anticoagulant*" or NOAC* or DOAC*).ti,ab.

or (apixaban or rivaroxaban or dabigatran or edoxaban).ti,ab.)

AND

((randomized controlled trial.pt.

or controlled clinical trial.pt.

or randomized.ab.

or placebo.ab.

or clinical trials as topic.sh.

or randomly.ab.

or trial.ti.)

not (exp animals/ not humans.sh.)))

[204 results]

#### **Database 4: Embase via Ovid**

| **Host** | Ovid (https://ovidsp.ovid.com); Embase 1974 to present |
| --- | --- |
| **Date searched** | 08 May 2026 |
| **Results** | 123 records |
| **RCT filter** | Emtree randomised controlled trial filter applied |
| **Note** | Conference abstracts excluded using a publication-type filter (.pt.). they provide insufficient detail for systematic data extraction in meta-analyses |

((exp atrial fibrillation/

or (atrial fibrillation or "non-valvular af").ti,ab.)

AND

(exp left atrial appendage closure/

or ((left atrial adj3 appendage) adj3

(closur* or occlusio* or exclusio* or ligatio*)).ti,ab.

or (laac or laao or "watchman flx" or "amplatzer amulet").ti,ab.)

AND

(exp direct oral anticoagulant/

or ("direct oral anticoagulant*" or noac* or doac*

or apixaban or rivaroxaban or dabigatran or edoxaban).ti,ab.)

AND

(exp randomized controlled trial/

or (randomized or randomly or placebo).ab.

or trial.ti.)

NOT (exp animal/ not exp human/))

NOT (conference abstract or conference review).pt.

[123 results]

#### **Database 5: ClinicalTrials.gov**

| **Host** | ClinicalTrials.gov Expert Search (https://clinicaltrials.gov/expert-search) |
| --- | --- |
| **Date searched** | 07 May 2026 |
| **Results** | 26 records |
| **RCT filter** | Not applied. Registry searched without study-type restriction to identify registered but unpublished or ongoing trials that may not yet appear in bibliographic databases |
| **Note** | Results screened for interventional RCTs comparing percutaneous LAAC against NOAC therapy. |

(AREA [Condition] EXPANSION [Concept] "Atrial Fibrillation")

AND

("Left Atrial Appendage Closure" OR LAAC OR LAAO OR WATCHMAN OR Amulet)

AND

("direct oral anticoagulant" OR NOAC OR DOAC

OR apixaban OR rivaroxaban OR dabigatran OR edoxaban)

AND AREA [StudyType] Interventional

[26 results]

#### **Search Summary**

| **Cochrane CENTRAL** | 129 records (07 May 2026) |
| --- | --- |
| **PubMed** | 222 records (08 May 2026) |
| **MEDLINE via Ovid** | 204 records (08 May 2026) |
| **Embase via Ovid** | 123 records (08 May 2026) |
| **ClinicalTrials.gov** | 26 records (07 May 2026) |
| **Total identified** | 704 records |
| **After deduplication** | 309 records |
| **Language restrictions** | None applied |
| **Date limits** | None applied |

#### Supplementary Table S1. Full-Text Excluded Studies with Reasons (PRISMA 2020)

| **#** | **Study / publication** | **Exclusion category** | **Specific reason for exclusion** |
| --- | --- | --- | --- |
| **Duplicate publications or registry entries (n = 8)** | | | |
| **1** | **CHAMPION-AF design / rationale paper**  Mansour M et al. Heart Rhythm. 2023. | **Duplicate publication** | Protocol paper; primary results subsequently published (NEJM 2026). No independent outcome data. |
| **2** | **CHAMPION-AF trial registry entry**  jRCT registry, 2021. | **Duplicate publication** | Registry entry superseded by published results. No outcome data. |
| **3** | **CLOSURE-AF design / rationale paper**  Landmesser U et al. Eur Heart J. 2026. | **Duplicate publication** | Protocol paper; primary results subsequently published (NEJM 2026). No independent outcome data. |
| **4** | **CLOSURE-AF EU trial registry entry**  EUCTR registry, 2017. | **Duplicate publication** | Early registry entry superseded by published results. |
| **5** | **OPTION design / rationale paper**  Wazni OM et al. Am Heart J. 2022. | **Duplicate publication** | Protocol paper; primary results subsequently published (NEJM 2025). No independent outcome data. |
| **6** | **PRAGUE-17 design / protocol paper**  Osmancik P et al. Am Heart J. 2017. | **Duplicate publication** | Protocol paper superseded by primary results publication (JACC 2020), which was selected as the included study. |
| **7** | **PRAGUE-17 4-year extended follow-up**  Osmancik P et al. J Am Coll Cardiol. 2022;79(1):1-14. | **Duplicate publication** | Extended follow-up of the same trial cohort. The 4-year data were used in this analysis from this publication; it is listed as a duplicate of the primary results paper to prevent double-counting. |
| **8** | **PRAGUE-17 ClinicalTrials.gov registry entry**  NCT02426944. ClinicalTrials.gov, 2015. | **Duplicate publication** | Registry entry only; results published separately as the included study. |
| **Sub-analyses and pilot studies not independently reporting pre-specified primary outcomes (n = 6)** | | | |
| **9** | **PRAGUE-17 heart failure biomarker sub-analysis**  Kocka V et al. Kardiol Pol. 2021. | **Sub-analysis / sub-study** | Biomarker sub-study (BNP, NT-proBNP). Does not report pre-specified clinical outcomes (stroke, embolism, mortality, or bleeding). Excluded to prevent double-counting. |
| **10** | **PRAGUE-17 nonprocedural bleeding sub-analysis (JACC Intv)**  Osmancik P et al. JACC Cardiovasc Interv. 2023.  Full title: Nonprocedural Bleeding After Left Atrial Appendage Closure Versus Direct Oral Anticoagulants: A Subanalysis of the Randomized PRAGUE-17 Trial. | **Sub-analysis / sub-study** | Sub-analysis of the PRAGUE-17 cohort focusing exclusively on nonprocedural bleeding outcomes. Does not independently report the primary composite endpoint or ischemic stroke. Excluded to prevent double-counting of PRAGUE-17 patients. |
| **11** | **OPTION sex-stratified sub-analysis**  OPTION investigators. 2023–2024. | **Sub-analysis / sub-study** | Secondary analysis of OPTION trial data stratified by sex. Does not independently report all pre-specified primary outcomes. Excluded to prevent double-counting of OPTION patients. |
| **12** | **OPTION AF-type sub-analysis**  OPTION investigators. 2023–2024. | **Sub-analysis / sub-study** | Secondary analysis of OPTION trial data by AF type (paroxysmal vs persistent). Does not independently report all pre-specified primary outcomes. Excluded to prevent double-counting. |
| **13** | **OPTION concomitant vs sequential sub-analysis**  Briceno DF et al. Heart Rhythm. 2025. doi:10.1016/j.hrthm.2025.04.029.  Full title: Comparison of Left Atrial Appendage Closure and Oral Anticoagulation After Catheter Ablation for Atrial Fibrillation: Concomitant and Sequential Cohorts of the OPTION Randomized Controlled Trial. | **Sub-analysis / sub-study** | Sub-analysis of the OPTION trial comparing same-day versus staged LAAC plus ablation procedures. Does not independently report the primary OPTION endpoints of stroke/embolism/CV death or non-procedural bleeding. Excluded to prevent double-counting of OPTION patients. |
| **14** | **Watchman versus rivaroxaban single-centre pilot RCT**  Registry: ChiCTR-PPR-15007044, 2015. | **Sub-analysis / sub-study** | Single-centre pilot study with no peer-reviewed results publication identified at time of search. Insufficient sample size and follow-up duration to satisfy inclusion criterion 4 (minimum 12 months, pre-specified outcomes). Classified as a pilot sub-study and excluded. |
| **No published outcome data, trial ongoing at time of search (n = 4)** | | | |
| **15** | **CATALYST trial design paper and registry entries**  Reddy VY et al. 2026 (design); NCT04226547 (registry). | **No outcome data (ongoing)** | Protocol paper and registry entries only. No outcome data available. Trial still enrolling (target n=2,650). Searched 8 May 2026. |
| **16** | **ELAPSE trial (formerly Occlusion-AF) design and registry**  Heidbuchel H et al. 2022 (design); NCT03067480 (registry). | **No outcome data (ongoing)** | Protocol paper and registry records only. No published outcome data identified. Trial enrollment ongoing at time of search. |
| **17** | **PROMOTE study protocol paper**  Shen L et al. BMJ Open. 2026. PMID 41698716. | **No outcome data (ongoing)** | Protocol paper only. Trial still enrolling (ChiCTR2000036538, target n=1,012). No outcome data available at time of search. |
| **18** | **LAAO versus NOAC in AF with recent PCI — registry entry**  NCT05353140. ClinicalTrials.gov, 2022. | **No outcome data (ongoing)** | Registry entry only. No published results. Trial enrolling AF patients with recent percutaneous coronary intervention. No outcome data available at time of search. |
| **Wrong population or design (n = 1)** | | | |
| **19** | **PVI-plus-LAAC versus PVI-plus-NOAC registry entry**  NCT06212674 / ChiCTR2000036538. Trial registry, 2022–2023. | **Wrong population / design** | Both trial arms mandate pulmonary vein isolation (PVI) as a co-intervention, precluding a pure LAAC-versus-NOAC comparison as specified in eligibility criterion 3. The ChiCTR entry is additionally a duplicate registry entry for the PROMOTE trial. |

#### * PRISMA 2020: Page MJ et al. BMJ. 2021;372:n71. doi:10.1136/bmj.n71. Records were managed and deduplicated using Rayyan (Ouzzani M et al. Syst Rev. 2016;5(1):210). Twenty records retrieved for full-text review were excluded. Exclusion reasons are categorised in accordance with PRISMA 2020 reporting requirements. Studies are listed in order of exclusion category. For studies with multiple records (e.g., design paper plus registry entry), each record is listed separately.

#### Abbreviations: AF = atrial fibrillation; LAAC = left atrial appendage closure; NOAC = non-vitamin K antagonist oral anticoagulant; PVI = pulmonary vein isolation; CV = cardiovascular; BNP = B-type natriuretic peptide; NT-proBNP = N-terminal pro-BNP; DXA = dual-energy X-ray absorptiometry.

Figure S1. Risk of Bias Assessment: Cochrane RoB 2 Traffic Light Plot

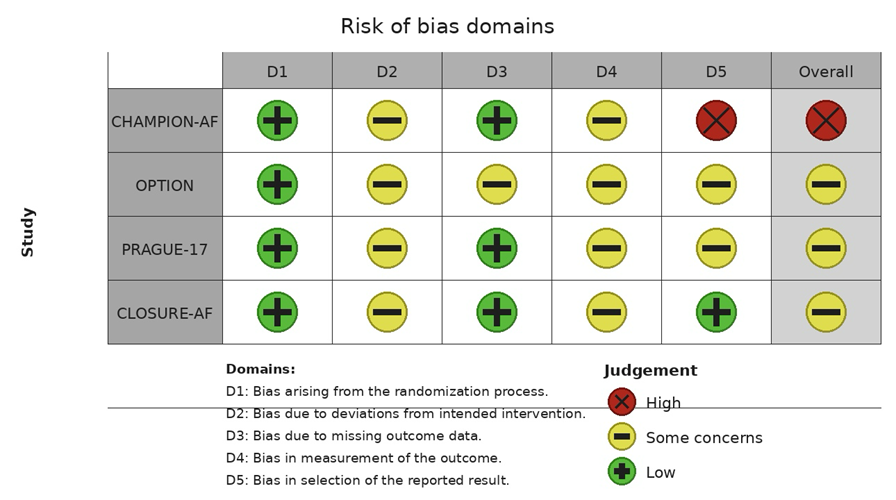

Risk of bias was assessed using the Cochrane RoB 2 tool across five domains for each included randomized controlled trial. While CHAMPION-AF was rated at high overall risk due to control-arm crossover and primary safety endpoint definitions, the remaining trials demonstrated some concerns reflecting their open-label designs, partially mitigated by blinded independent Clinical Events Committees. NOAC = non-vitamin K antagonist oral anticoagulant; RCT = randomized controlled trial; RoB = risk of bias.

### Supplemental Results

**Supplementary Table S2. Exploratory Pooled Estimates: Primary Efficacy Composite and Non-Procedural Bleeding**

| **Trial** | **Efficacy Composite  HR/sHR/IRR (95% CI)** | **I²** | **Bleeding  HR/sHR/IRR (95% CI)** | **I²** |
| --- | --- | --- | --- | --- |
| **CHAMPION-AF** | 1.20 (0.87–1.66) | — | 0.55 (0.45–0.67) | — |
| **OPTION** | 0.91 (0.59–1.39) | — | 0.44 (0.33–0.59) | — |
| **PRAGUE-17** | 0.81 (0.56–1.18) | — | 0.55 (0.31–0.97) | — |
| **CLOSURE-AF** | 1.27 (1.00–1.60) | — | 0.89 (0.61–1.28) | — |
| **Pooled (exploratory)¶** | **1.07 (0.86–1.33)** | **41%** | **0.58 (0.44–0.76)** | **65%** |

*¶ Pooled efficacy and bleeding estimates are exploratory only. HR = Cox hazard ratio; sHR = Fine-Gray subdistribution HR (PRAGUE-17); IRR = Poisson incidence rate ratio (CLOSURE-AF). Pooling on the log-hazard scale is justified under rare-event approximation (cumulative efficacy event rates <12%; bleeding rates 8.5–19.0% approach threshold — interpret with caution). These figures are numerically consistent with the narrative synthesis in the main manuscript and serve as internal consistency checks only.*

**Section 1. Exploratory Pooled Forest Plots (Reference Only)**

The four included trials used different primary statistical frameworks: CHAMPION-AF and OPTION used standard Cox hazard ratios; PRAGUE-17 used Fine-Gray subdistribution hazard ratios; CLOSURE-AF required restricted mean survival time for bleeding due to proportional hazards violation and Poisson incidence rate ratios for other outcomes. Pooling these measures on the log-hazard scale is not the primary analytical approach but is provided as an exploratory internal consistency check. The resulting pooled estimates are directionally and numerically consistent with the narrative synthesis presented in the main manuscript, confirming that the clinical conclusions are not sensitive to the choice of analytical framework.

**Figure S2. Primary Efficacy Composite: All Four Trials (Exploratory)**

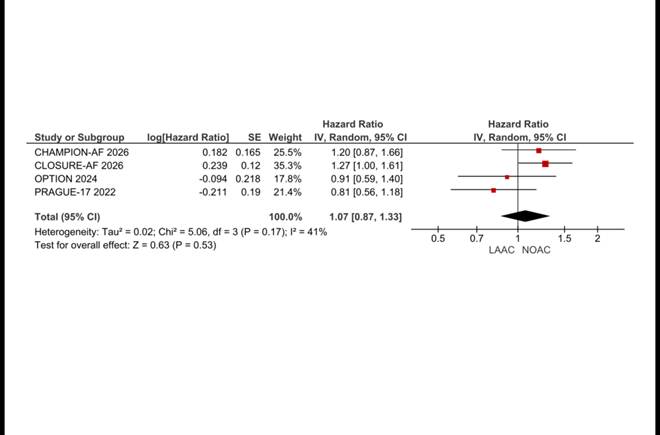

*Exploratory pooled estimate: HR 1.07 (95% CI 0.87–1.33), I²=41%, P=0.53. Provided for reference only. CHAMPION-AF and OPTION report Cox HRs; PRAGUE-17 reports Fine-Gray sHR; CLOSURE-AF reports Poisson IRR. Pooling is exploratory under rare-event approximation. HR >1 favours NOAC; HR <1 favours LAAC.*

**Figure S3. Non-Procedural Bleeding: All Four Trials (Exploratory)**

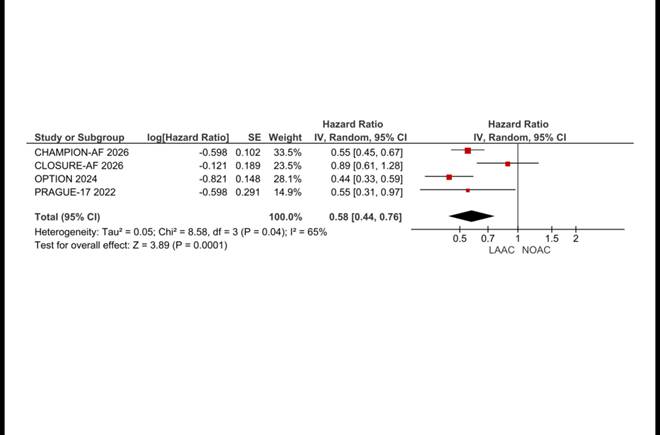

*Exploratory pooled estimate: HR 0.58 (95% CI 0.44–0.76), I²=65%, P=0.0001. Provided for reference only. Cumulative bleeding incidences of 8.5–19.0% exceed the rare-event threshold for valid pooling of different effect measures. CLOSURE-AF violated proportional hazards for bleeding; its IRR is therefore not directly comparable to the Cox HRs from the other three trials. HR <1 favours LAAC.*

**Section 2. Leave-One-Out Sensitivity Analyses**

Pre-specified leave-one-out analyses were conducted by sequentially excluding each trial for all three outcomes. These analyses identify which trials drive heterogeneity and test robustness of the primary findings. Figures S4–S11 present the corresponding leave-one-out analyses for the exploratory efficacy composite and bleeding outcomes. For ischemic stroke leave-one-out figures are presented in Supplementary Figures S12–S15.

***Primary Efficacy Composite: Leave-One-Out Analyses (Exploratory)***

**Figure S4. Primary Efficacy Composite: Excluding CLOSURE-AF**

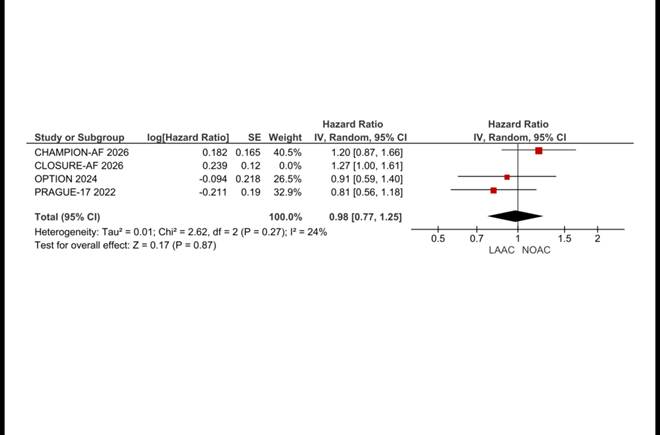

*Excl. CLOSURE-AF: HR 0.98 (95% CI 0.77–1.25), I²=24%, P=0.87. Exclusion of CLOSURE-AF reduces heterogeneity from 41% to 24% and shifts the estimate to 0.98, consistent with noninferiority across the three moderate-risk trials. Confirms CLOSURE-AF as the principal driver of heterogeneity for this outcome.*

**Figure S5. Primary Efficacy Composite: Excluding CHAMPION-AF**

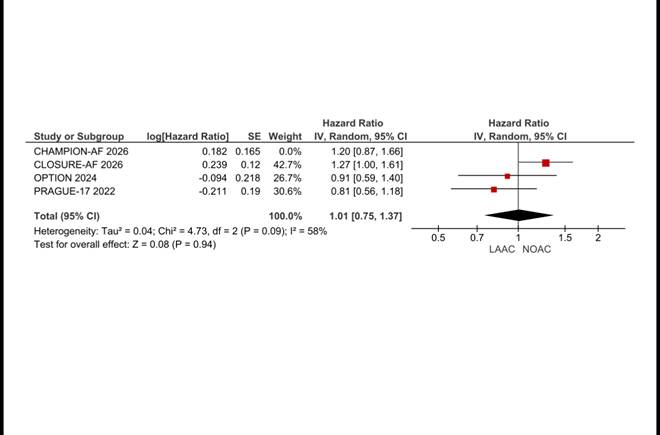

*Excl. CHAMPION-AF: HR 1.01 (95% CI 0.75–1.37), I²=58%, P=0.94. Heterogeneity remains moderate after CHAMPION-AF exclusion, driven by the contrast between CLOSURE-AF and the remaining moderate-risk trials.*

**Figure S6. Primary Efficacy Composite: Excluding OPTION**

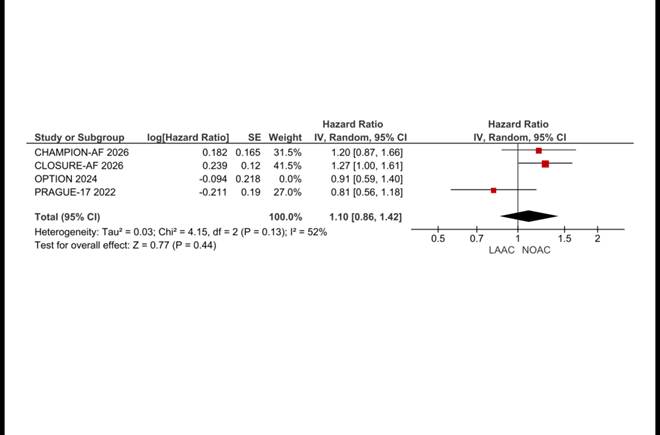

*Excl. OPTION: HR 1.10 (95% CI 0.86–1.42), I²=52%, P=0.44.*

**Figure S7. Primary Efficacy Composite: Excluding PRAGUE-17**

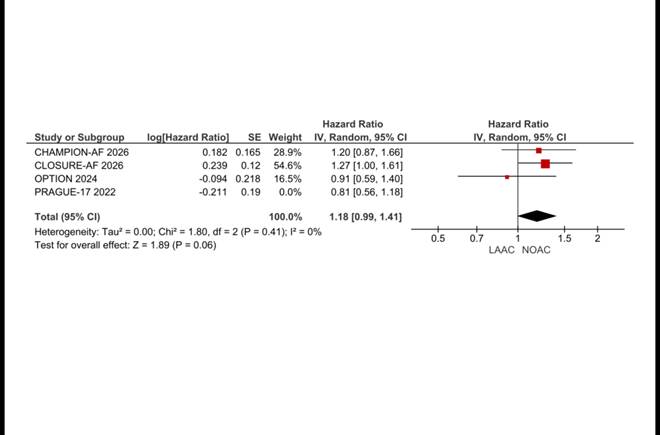

*Excl. PRAGUE-17: HR 1.18 (95% CI 0.99–1.41), I²=0%, P=0.06. Exclusion of PRAGUE-17 resolves heterogeneity completely (I²=0%), confirming that its Fine-Gray sHR is the source of residual variance once CLOSURE-AF is excluded.*

***Non-Procedural Bleeding: Leave-One-Out Analyses (Exploratory)***

**Figure S8. Non-Procedural Bleeding: Excluding CLOSURE-AF**

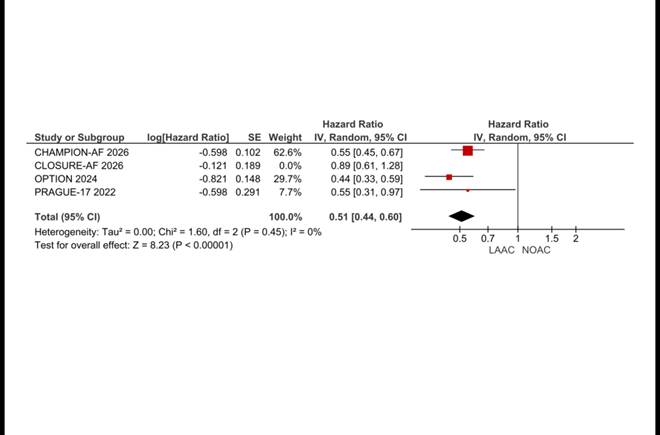

*Excl. CLOSURE-AF: HR 0.51 (95% CI 0.44–0.60), I²=0%, P<0.00001. Complete resolution of heterogeneity on CLOSURE-AF exclusion confirms it as the sole driver of between-trial variance for bleeding. The three moderate-risk trials (CHAMPION-AF, OPTION, PRAGUE-17) are highly consistent in demonstrating a 45–56% bleeding reduction with LAAC.*

**Figure S9. Non-Procedural Bleeding: Excluding CHAMPION-AF**

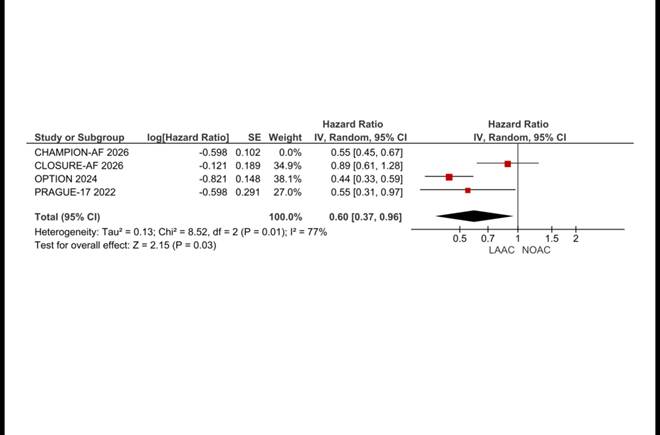

*Excl. CHAMPION-AF: HR 0.60 (95% CI 0.37–0.96), I²=77%, P=0.03. High residual heterogeneity confirms CLOSURE-AF remains the primary driver even after CHAMPION-AF exclusion.*

**Figure S10. Non-Procedural Bleeding: Excluding OPTION**

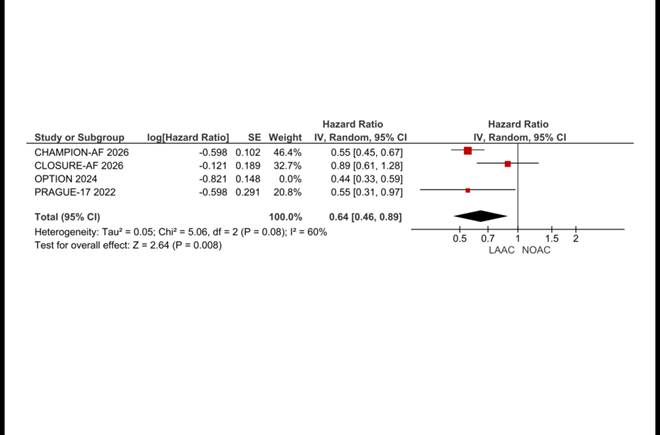

*Excl. OPTION: HR 0.64 (95% CI 0.46–0.89), I²=60%, P=0.008.*

**Figure S11. Non-Procedural Bleeding: Excluding PRAGUE-17**

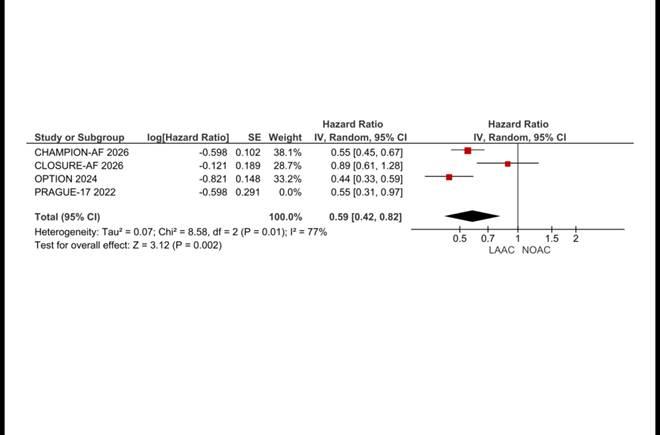

*Excl. PRAGUE-17: HR 0.59 (95% CI 0.42–0.82), I²=77%, P=0.002. High heterogeneity persists, confirming CLOSURE-AF as the dominant source of variance regardless of which other trial is excluded.*

#### ***Ischemic Stroke: Leave-One-Out Sensitivity Analyses***

**Figure S12. Leave-One-Out: Excluding CHAMPION-AF**

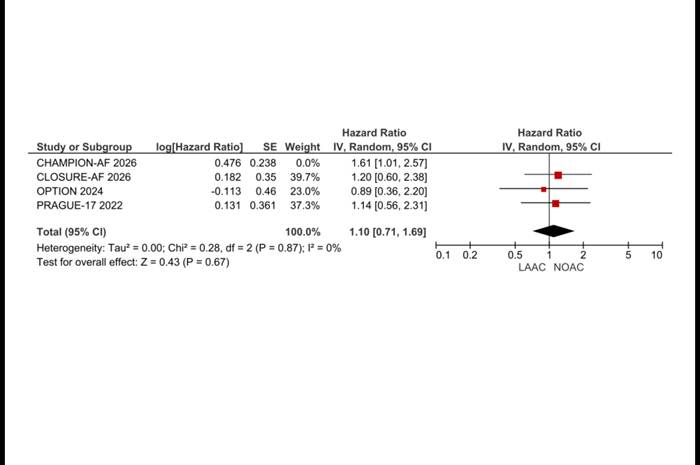

*Pooled HR 1.10 (95% CI 0.71–1.69, P=0.67, I²=0%). The substantial attenuation on CHAMPION-AF exclusion identifies it as the principal driver of the directional stroke signal.*

**Figure S13. Leave-One-Out: Excluding CLOSURE-AF**

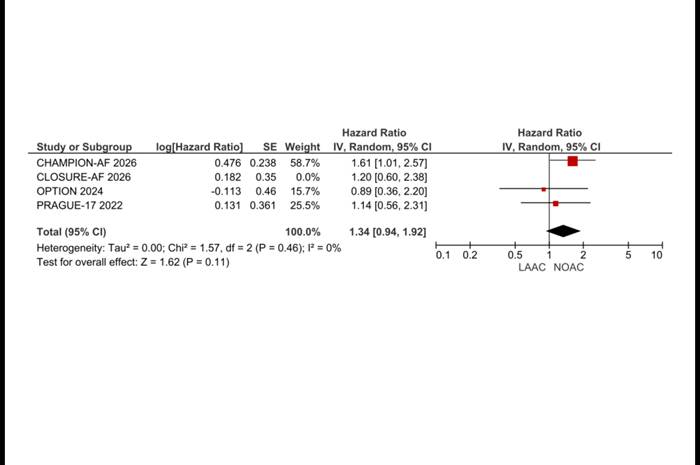

*Pooled HR 1.34 (95% CI 0.94–1.92, P=0.11, I²=0%).*

**Figure S14. Leave-One-Out: Excluding OPTION**

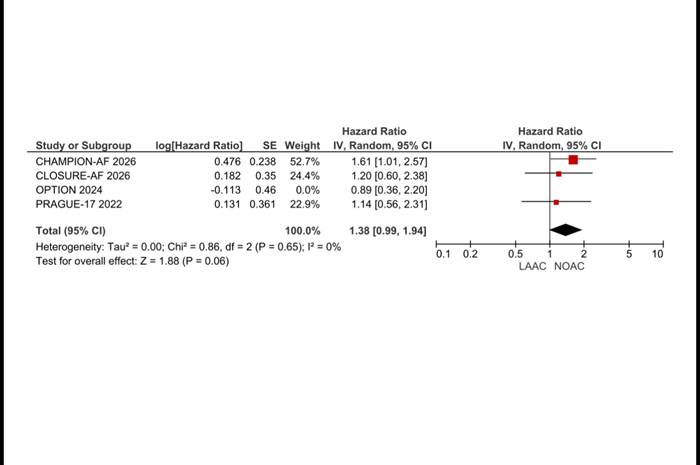

*Pooled HR 1.38 (95% CI 0.99–1.94, P=0.06, I²=0%).*

**Figure S15. Leave-One-Out: Excluding PRAGUE-17**

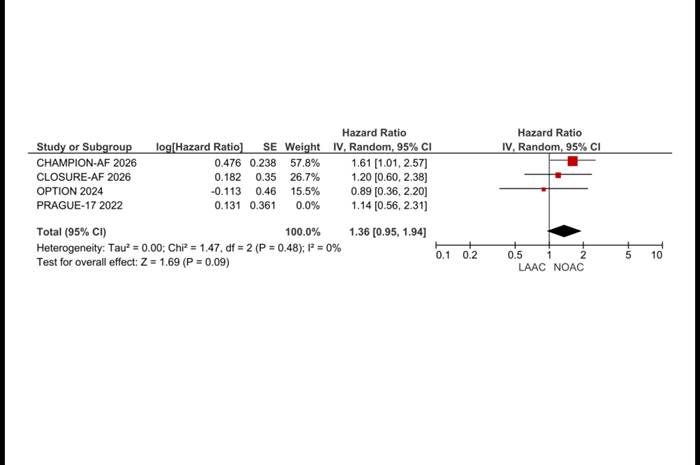

*Pooled HR 1.36 (95% CI 0.95–1.94, P=0.09, I²=0%).*
